# Estimating and Testing an Index of Bias Attributable to Composite Outcomes in Comparative Studies

**DOI:** 10.1101/2020.02.13.20020966

**Authors:** Fredi Alexander Diaz-Quijano

## Abstract

**Objective:** To develop an index to evaluate the bias attributable to composite outcomes (BACO) in comparative clinical studies.

**Study design and Setting:** I defined the BACO index as the ratio of the logarithm of the association measure (e.g., relative risk) of the composite outcome to that of its most relevant component endpoint (e.g., mortality). Methods to calculate the confidence intervals and test the null hypotheses (BACO index = 1) were described and applied in systematically selected clinical trials. Two other preselected trials were included as “positive controls” for being examples of primary composite outcomes disregarded due to inconsistency with the treatment effect on mortality.

**Results:** The BACO index values different from one were classified according to whether the use of composite outcomes overestimated (BACO index >1), underestimated (BACO index between zero and <1), or inverted (BACO index <0) the association between exposure and prognosis. In three of 23 systematically selected trials and the two positive controls, the BACO indices were significantly lower than one (p <0.005).

**Conclusion:** BACO index can warn that the composite outcome association is stronger, weaker, or even opposite than that of its most critical component.

What is new?
- BACO index is the ratio of the logarithm of the association measure (e.g., RR) of the composite outcome to the logarithm of the association measure of the component endpoint that represents the study target (e.g., mortality).
- In comparative studies, the BACO index can be used to evaluate the correspondence between the effect on a composite outcome and that on its most critical component.
- This index could help to preset rules to make decisions for interpretation of clinical studies.
- A significant BACO should lead to the caution that the association of the composite is stronger (BACO index >1), weaker (BACO index between zero and <1), or even opposite (BACO index <0) than that of its most critical component.

## 1. Introduction

Composite outcomes, particularly those that define an event when at least one of a group of component endpoints occurs, are increasingly used in clinical research [1–3]. In the last two decades, publications using terms referring to composite outcomes increased progressively in PubMed, from less than one hundred per year, before 2004, to more than one thousand per year since 2018. About a third of these publications are associated with the terms “clinical” and “trial” (Figure 1). These figures highlight the importance of this kind of outcome to evaluate interventions in clinical practice.

**Figure 1.**
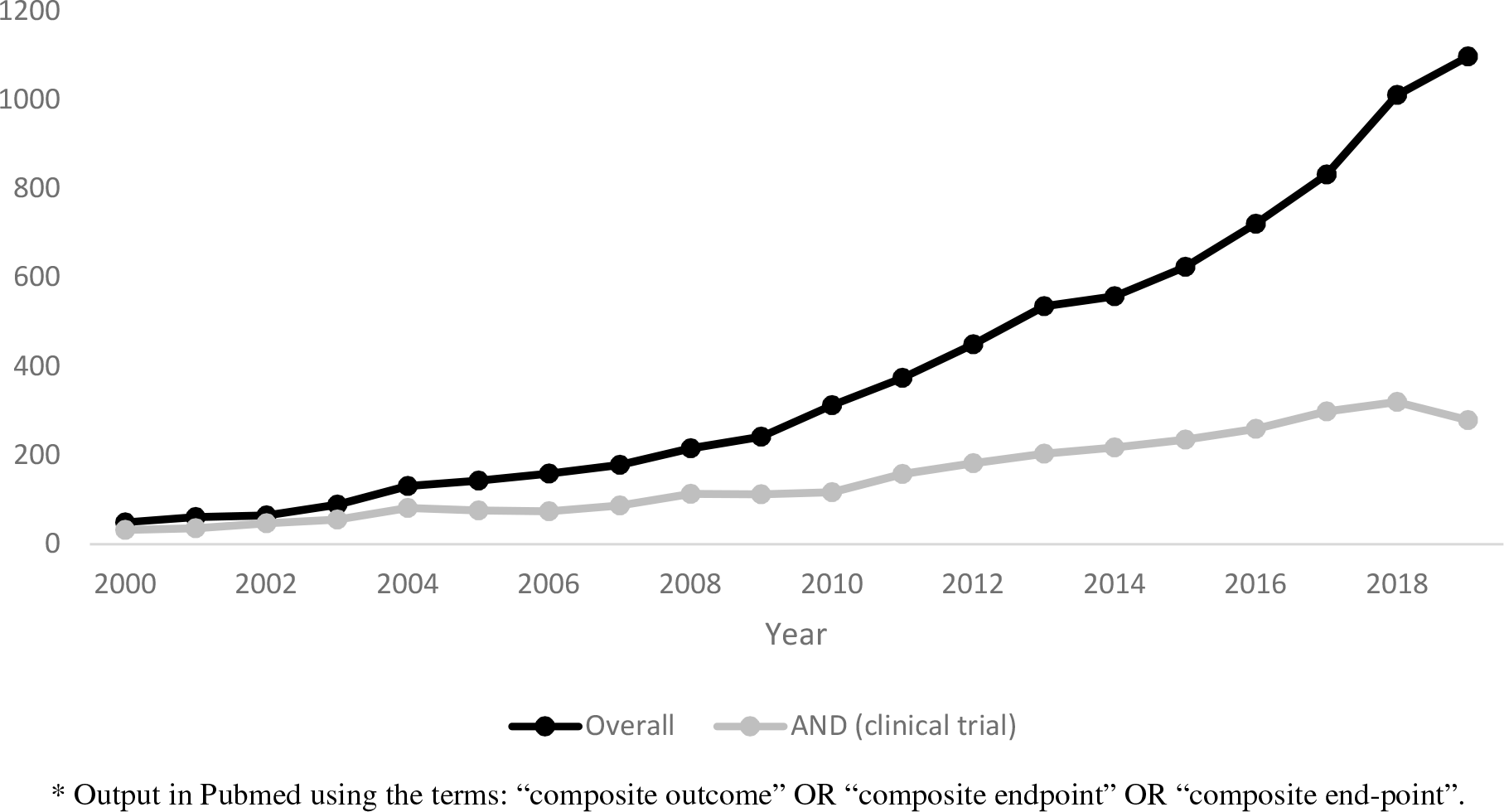
Annual counts of publications using terms related to Composite Outcome.*

The use of a composite outcome can have different methodological purposes [3,4]. When several potentially eligible endpoints exist, a composite outcome can avoid the need to choose a simple one and prevent problems associated with multiple comparisons [1,4]. Moreover, it can be a way to deal with competing risks [5]; or deliberately integrate events of different nature, such as indicators of the effectiveness and safety of an intervention [6].

In many other instances, composite outcomes are used to represent severity more broadly than with a single event (e.g., death). This practice facilitates a higher number of events for analysis, which increases the power of the study, reduces its costs, and provides a faster response to a research question [1,2,4]. However, the association measures are often misinterpreted as the effect of exposure on each of the elements of the composite outcome [2,7]. This is problematic when the overall effect on the composite outcome does not follow the same direction as its most critical components.

In some cases, effects on primary composite outcomes have been ruled out because they differ greatly from those on mortality [8–10]. This may occur because the composite outcome includes events generated by different mechanisms. For example, some components may be directly related to severity (heart attacks, death), while others may be more influenced by medical decisions and resource availability (hospitalization, catheterization). Therefore, some component endpoints can introduce bias that affects the estimation of the overall effect.

Despite their importance, little guidance is available on how composite outcomes should be interpreted, especially in situations of varied direction in the association across the event subtypes [4,6,11]. Moreover, there is a lack of statistical tools to support the decision to accept or rule out the use of the composite outcome. Furthermore, because composite outcomes are inherently correlated with their components [7], comparative estimates to quantify and test biases associated with the use of this type of endpoint are challenging to make.

In this paper, I proposed an index to evaluate the bias attributable to the composite outcome (BACO), which is simply the ratio of the logarithm of the association measure of the composite outcome to that of the component endpoint that represents the study target (e.g., mortality). For this purpose, I described how to calculate confidence intervals and perform hypothesis tests. Then, I applied these procedures in a group of clinical trials recently published in major medical journals.

## 2. Methods

### 2.1. BACO Index

I aimed to compare the association measure of a composite outcome with that obtained with a component endpoint, which indisputably represents the study target. Here, I chose the any-cause mortality as the target [12], and defined the BACO index as follows:

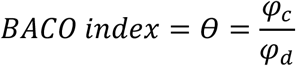

Where *φ_c_* and *φ_d_* are the natural logarithms (*ln*) of ratio-based association measures for a composite outcome and death, respectively. In this paper, I mean relative risk (RR) as the association measure; however, the concept could be applied to other measures (e.g., hazard ratio or incidence rate ratio).

A BACO index equal to one indicates that there is no bias attributable to the use of a composite outcome. A BACO index higher than one indicates that the association with the composite outcome is stronger than that with death. A value between zero but less than one suggests that the association with the composite outcome is biased towards nullity. On the other hand, a negative value would result when the bias leads to an inversion of the association.

These interpretations can be applied regardless of the reference group in the comparative study. Because if the comparison groups were inverted, the signs of both the numerator and denominator would also be inverted.

Since the BACO index is a ratio between two correlated variables, calculating its variance, *υ(θ*), requires considering the covariance between *φ_c_* and *φ_d_*. Specifically, by applying the Taylor series estimator [13], we have that

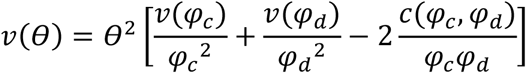

Where *υ*(*φ_c_*) and *υ*(*φ_d_*) are the variances of *φ_c_* and *φ_d_*, respectively, and *c*(*φ_c,_ φ_d_*), is their covariance. This covariance is usually calculated through complex matrix operations [13,14]. However, in the case of the BACO index, we analyze effects on two outcomes, one of which is completely embedded within the other. Therefore, *φ_c_* can be disaggregated as a sum of two effects: on death and on the other component endpoints, namely, *φ_c_*.= *φ_d_* + *φ_c_*_−_*_d_*. As detailed in the Appendix A, with this approach, I deduced that *c*(*φ_c,_ φ_d_*) = *υ*(*φ_c_*). Consequently, the formula of *υ*(*θ*) can be simplified to:

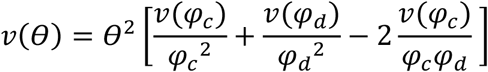

A 95% confidence interval for the BACO index is

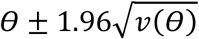

And the statistic with normal distribution to test the null hypothesis (BACO index = 1) is

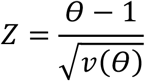

Thus, the BACO index can be estimated and tested directly from contingency tables (Appendix A). Alternatively, we can combine regression parameters, which may be useful to estimate a BACO index from adjusted association measures. For example, the *ln*(RR) can be estimated as the coefficient of a Poisson regression, which is an alternative to avoid the problems of convergence of the Log-Binomial regression [15]. After, we can combine the results into one parameter vector and simultaneous (co)variance matrix of the sandwich/robust type, which is appropriate even if the estimates were obtained with the same or overlapping data [16]. Then, using the non-linear combination of parameters, the BACO indices can be calculated as the ratio of the regression coefficient of the composite outcome to that of mortality (see example in Appendix B). These procedures lead to results virtually identical to those obtained directly from contingency tables (Appendix C).

### 2.2. Estimation with simulated data

To illustrate different results of the BACO index, I simulated a comparative study in which a group of a thousand people was exposed to an experimental intervention and presented mortality of 4.8% during the follow-up. This group was compared with a reference group of another thousand people, which showed an 8% mortality. Consequently, the RR for death was 0.6 (4.8%/8%), in other words, the intervention had an efficacy of 40% for reducing mortality.

Now consider four composite outcomes, all of which included death within their components. The first composite outcome maintained the same RR as observed for mortality. The second outcome led to an overestimation of the treatment effect with a RR of 0.3 (efficacy of 70%). On the contrary, the third outcome led to an underestimation of the effect with a RR of 0.8 (efficacy of 20%). Finally, the fourth composite outcome led to an inversion of the association measure, suggesting that the intervention duplicates the risk of the event (Table 1).

**Table 1.**
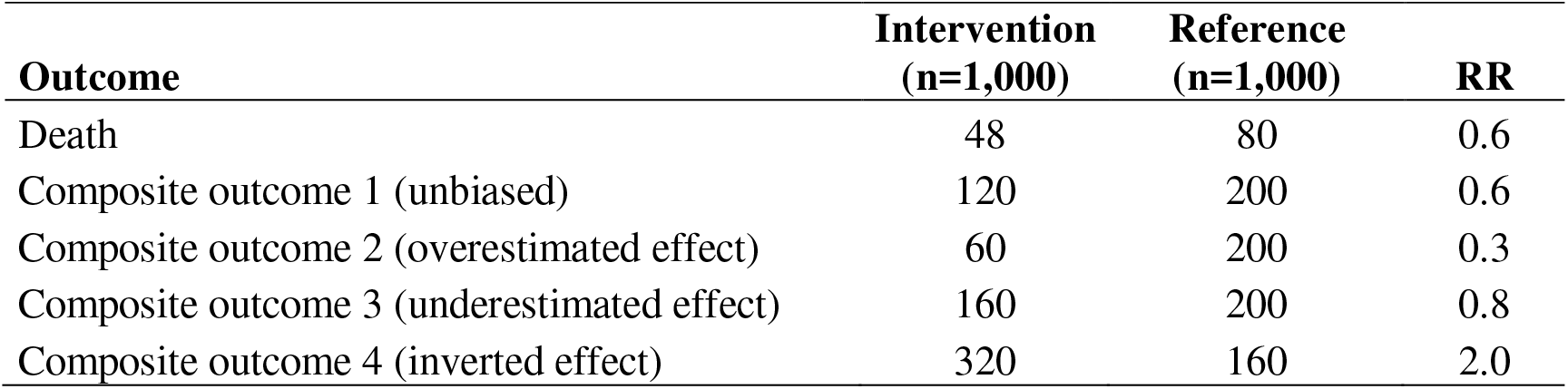
Frequency of simulated outcomes in two hypothetic groups.

| Outcome | Intervention<br>(n=1,000) | Reference<br>(n=1,000) | RR |
| --- | --- | --- | --- |
| Death | 48 | 80 | 0.6 |
| Composite outcome 1 (unbiased) | 120 | 200 | 0.6 |
| Composite outcome 2 (overestimated effect) | 60 | 200 | 0.3 |
| Composite outcome 3 (underestimated effect) | 160 | 200 | 0.8 |
| Composite outcome 4 (inverted effect) | 320 | 160 | 2.0 |

Figure 2 illustrates the point estimates and the corresponding 95% confidence intervals (95% CI). For the first composite outcome (unbiased), the BACO index was equal to one (95% CI: 0.46 to 1.54). For the second composite outcome, which overestimated the effect of the intervention on prognosis, the BACO index was 2.36 (95% CI: 1.14 to 3.58). The third outcome, which underestimated the effect, presented a BACO index of 0.44 (95% CI: 0.11 to 0.76). For the last simulated composite outcome, which had an inverted association measure, the BACO index was negative: -1.36 (95% CI: -2.48 to -0.24).

**Fig. 2.**
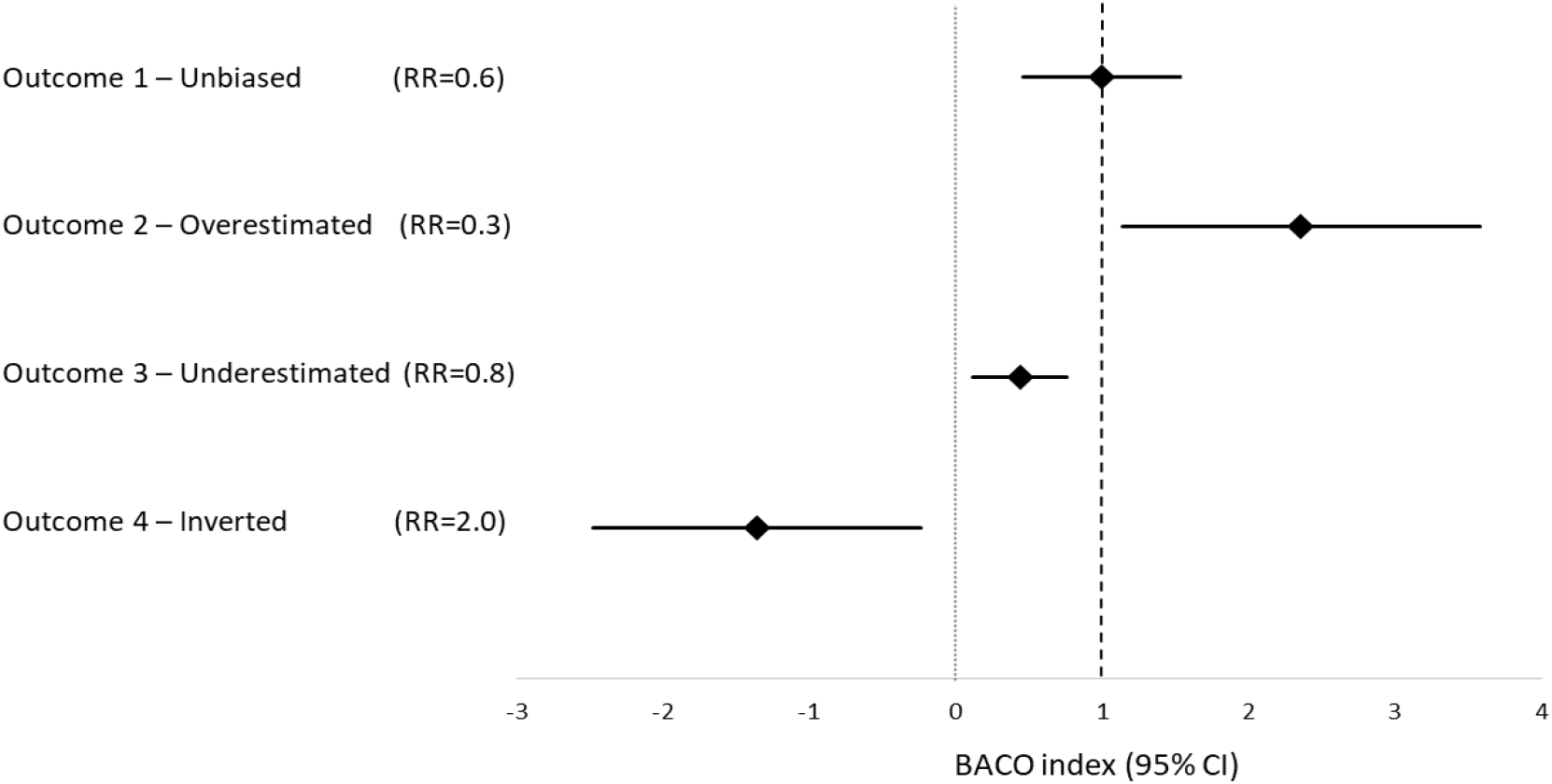
BACO indices of four simulated composite outcomes.

### 2.3. Application

I searched PubMed to identify 2-group, parallel-design clinical trials published in the journal groups of JAMA, NEJM, and Lancet in 2019 (updated on 07/01/2020), using the following word combination: composite primary (endpoint OR outcome OR (“end-point”)) (mortality OR death) (randomised OR randomized) (trial) (JAMA OR NEJM OR Lancet). Next, I selected those studies whose primary outcome was a composite, binary, and included all-cause mortality within their components. Secondary subgroup analyses and studies with five or fewer fatal events were excluded. A study was also excluded whose outcome results were mainly based on imputations because it presented substantial losses during the follow-up [17].

I reviewed each article and built a database to reproduce individual information for the variables of intervention, composite outcome, and death. When necessary, I contacted the corresponding author to ask for data not provided in the article. For each trial, I calculated the RR for both the composite outcome (RR_c_) and death from any cause (RR_d_), using the corresponding intervention as the independent variable. After, I estimated the BACO index as a ratio of regression coefficients and performed Wald-type tests, based on the delta method, for the null hypothesis that the BACO index is equal to one. These analyses were conducted using Stata software (version 15.0, Stata Corp LP, College Station, TX, USA), and the commands employed are described in the Appendix B.

This work intended to reproduce the independent application of the BACO index in each of the studies. Therefore, I did not consider the number of trials to adjust the level of significance. However, I preset 0.005 as a level to define a statistically significant BACO [18]. A higher level (e.g., 0.05) was not chosen because I assumed that the composite outcomes were purposefully defined to be consistent with mortality, thus expecting a low pre-test probability that the BACO index is different from one. However, I used the term “suggestive” for p values between 0.005 and 0.05 [19].

Besides the selected studies from 2019, I also analyzed the clinical trials CAPRICORN and EXPEDITION [20,21]. CAPRICORN investigated carvedilol in patients with left-ventricular dysfunction after acute myocardial infarction [20]; and EXPEDITION evaluated intravenous caripode in high-risk coronary artery bypass graft surgery patients [21]. These two studies were included as “positive controls” because they represented examples of primary composite outcomes that were disregarded for being inconsistent with the treatment effect on mortality [9,10].

## 3. Results

From 82 articles, I selected 23 clinical trials, most of which were about cardiovascular diseases. Besides mortality, the composite outcomes integrated diverse components, often including cardio-cerebrovascular events, such as myocardial infarction or stroke, and other related to the use of health services, such as hospital admission and vascular interventions. The sample sizes of these trials ranged from 240 to 536,233. While the number of patients evolving composite outcomes ranged from 31 to 4,067, the number of deaths ranged from six to 1,140 (Table 2).

**Table 2.**
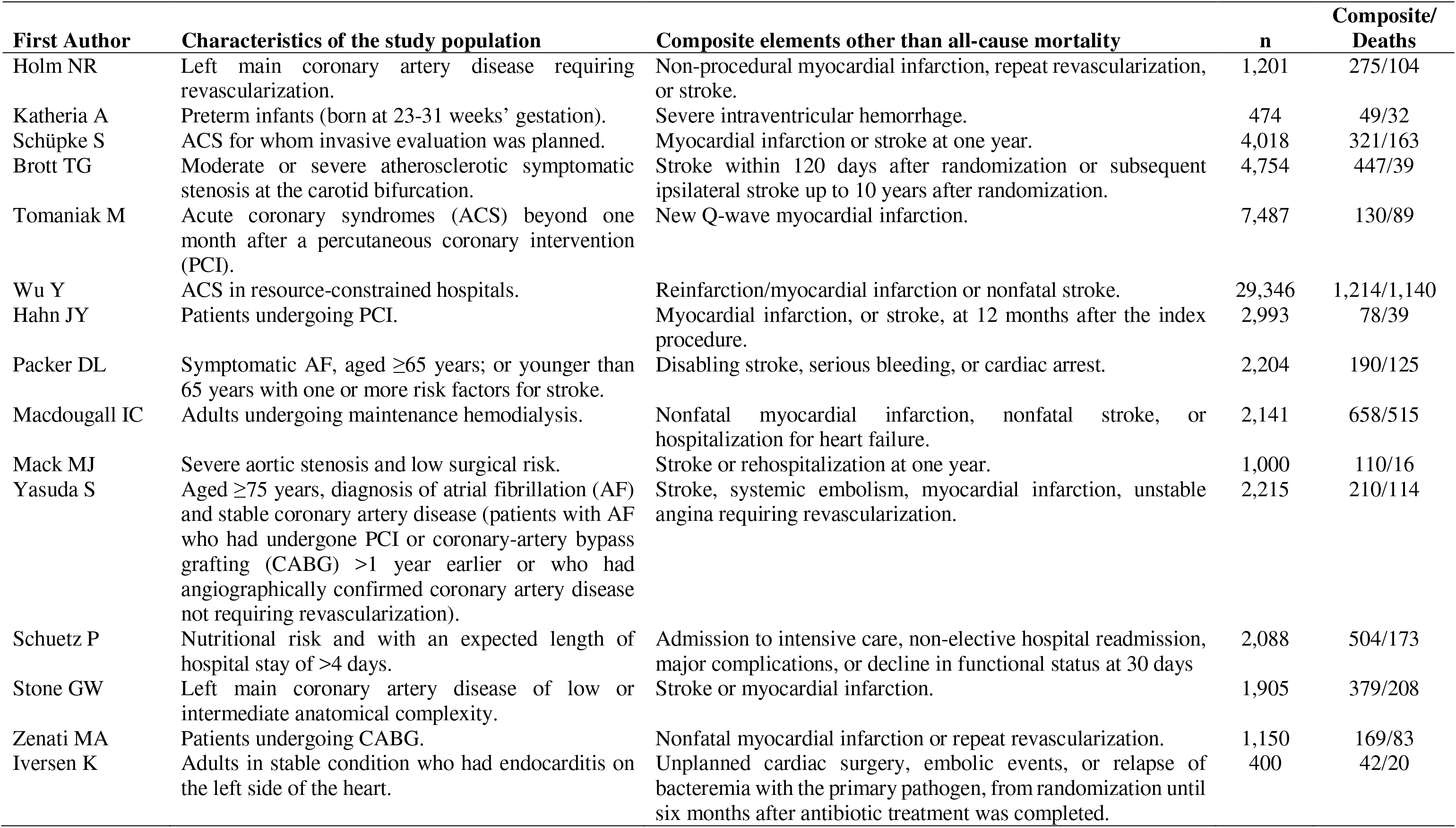
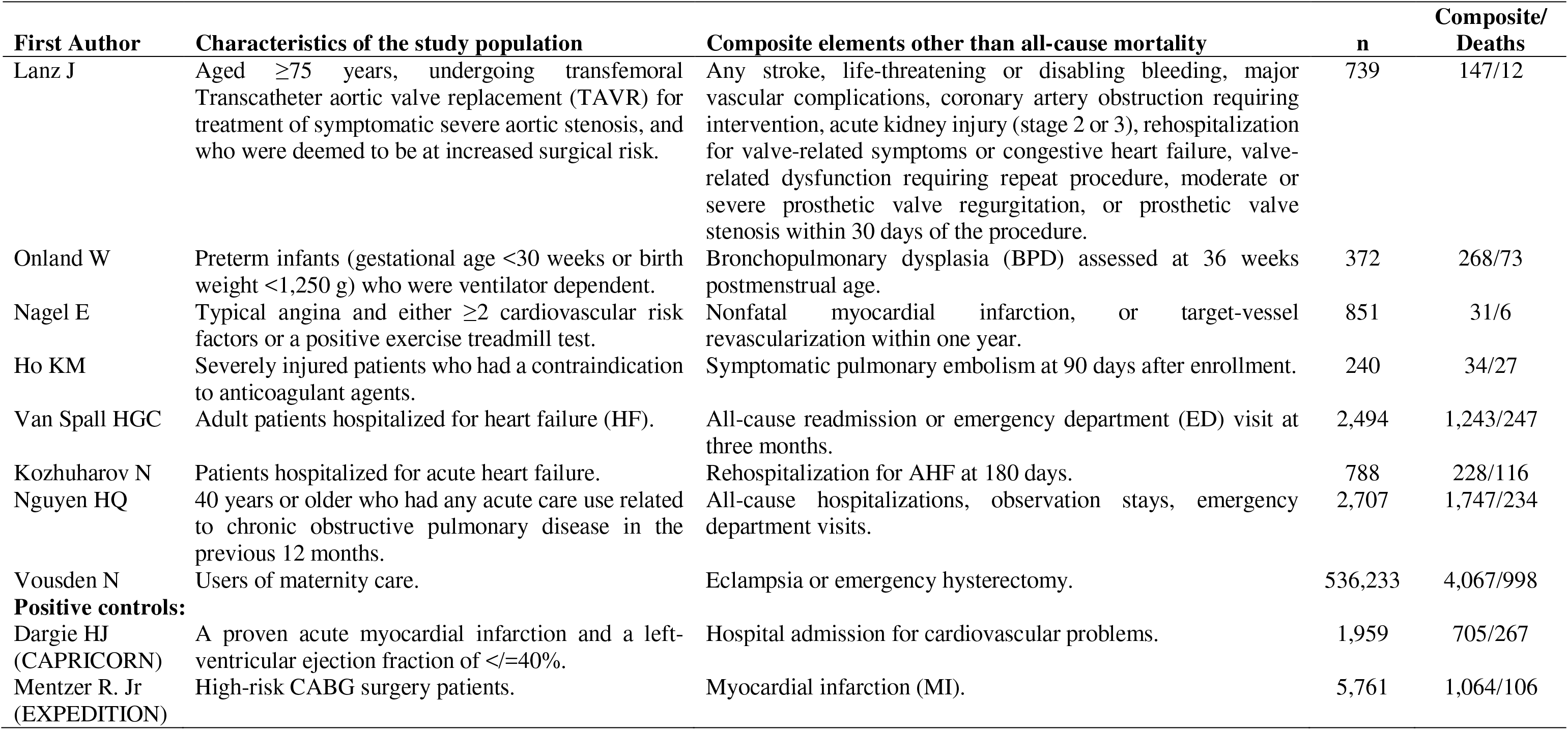
General description of the study population and outcomes of clinical trials selected.

In six studies [22–27], the RRs of the composite outcomes were further from the null value than the corresponding RRs of death (Table 3). Consequently, their BACO indices were greater (although not statistically different) than one. One study had the same association measure for both the composite outcome and any-cause death (BACO index = 1) [28].

**Table 3.** Relative risks of composite and death, and BACO index in clinical trials.

| First author | RR<sub>c</sub> (95% CI) | RR<sub>d</sub> (95% CI) | BACO index (95% CI) | p value* |
| --- | --- | --- | --- | --- |
| Holm NR | 1.51 (1.22 to 1.87) | 1.09 (0.75 to 1.57) | 4.85 (-14.7 to 24.4) | 0.70 |
| Katheria A | 1.46 (0.85 to 2.51) | 1.14 (0.58 to 2.23) | 2.84 (-8.44 to 14.1) | 0.75 |
| Schüpke S | 1.34 (1.08 to 1.66) | 1.23 (0.91 to 1.66) | 1.41 (-0.12 to 2.95) | 0.60 |
| Brott TG | 0.71 (0.59 to 0.85) | 0.78 (0.42 to 1.47) | 1.41 (-2.08 to 4.9) | 0.82 |
| Tomaniak M | 0.73 (0.52 to 1.03) | 0.74 (0.49 to 1.13) | 1.05 (0.22 to 1.89) | 0.90 |
| Wu Y | 0.87 (0.78 to 0.97) | 0.87 (0.78 to 0.97) | 1 (0.8 to 1.21) | 0.99 |
| Hahn JY | 1.17 (0.75 to 1.81) | 1.17 (0.63 to 2.19) | 1 (-1.85 to 3.85) | 1 |
| Packer DL | 0.87 (0.66 to 1.14) | 0.86 (0.61 to 1.2) | 0.89 (-0.31 to 2.08) | 0.85 |
| Macdougall IC | 0.91 (0.8 to 1.03) | 0.88 (0.75 to 1.02) | 0.74 (0.22 to 1.26) | 0.32 |
| Mack MJ | 0.61 (0.04 to 0.88) | 0.45 (0.16 to 1.28) | 0.62 (-0.16 to 1.4) | 0.34 |
| Yasuda S | 0.74 (0.57 to 0.96) | 0.56 (0.39 to 0.82) | 0.53 (0.21 to 0.86) | 0.0048 |
| Schuetz P | 0.84 (0.72 to 0.98) | 0.72 (0.54 to 0.96) | 0.52 (0.07 to 0.98) | 0.04 |
| Stone GW | 1.16 (0.97 to 1.4) | 1.35 (1.04 to 1.75) | 0.51 (0.07 to 0.94) | 0.03 |
| Zenati MA | 1.12 (0.84 to 1.48) | 1.25 (0.82 to 1.89) | 0.5 (-0.45 to 1.44) | 0.30 |
| Iversen K | 1.35 (0.75 to 2.4) | 1.88 (0.76 to 4.6) | 0.47 (-0.24 to 1.18) | 0.15 |
| Lanz J | 1.43 (1.06 to 1.92) | 2.96 (0.81 to 10.8) | 0.33 (-0.1 to 0.76) | 0.002 |
| Onland W | 0.95 (0.84 to 1.08) | 0.65 (0.42 to 0.99) | 0.11 (-0.17 to 0.39) | <0.001 |
| Nagel E | 0.96 (0.48 to 1.91) | 2.04 (0.38 to 11.1) | -0.06 (-1.1 to 0.98) | 0.04 |
| Ho KM | 0.97 (0.52 to 1.8) | 1.41 (0.68 to 2.9) | -0.1 (-2.11 to 1.91) | 0.28 |
| Van Spall HGC | 0.98 (0.91 to 1.06) | 1.03 (0.81 to 1.3) | -0.63 (-7.64 to 6.38) | 0.65 |
| Kozhuharov N | 1.1 (0.88 to 1.37) | 0.94 (0.67 to 1.31) | -1.48 (-12 to 9.05) | 0.64 |
| Nguyen HQ | 1.02 (0.96 to 1.07) | 0.99 (0.78 to 1.27) | -2.27 (-88.3 to 83.7) | 0.94 |
| Vousden N | 0.92 (0.86 to 0.97) | 1.04 (0.92 to 1.18) | -2.35 (-11.2 to 6.51) | 0.46 |
| **Positive Controls:** |  |  |  |  |
| Dargie HJ (CAPRICORN) | 0.94 (0.84 to 1.06) | 0.78 (0.62 to 0.97) | 0.24 (-0.15 to 0.64) | <0.001 |
| Mentzer R. Jr (EXPEDITION) | 0.82 (0.73 to 0.91) | 1.53 (1.04 to 2.26) | -0.48 (-1.03 to 0.08) | <0.001 |

\* p-value for the null hypothesis of the BACO index is equal to one.
RR<sub>c</sub>: Relative Risk for the composite outcome; RR<sub>d</sub>: Relative Risk for any-cause death; BACO index: $\ln(RR_c)/\ln(RR_d)$

BACO index was lower than one in the other 16 studies [29,30,39–44,31–38], and the BACO was statistically significant in three of them. These included the study by Yasuda et al., in which a monotherapy with rivaroxaban was compared to a combination therapy with rivaroxaban plus a single antiplatelet agent [32]. The apparent effect of the monotherapy on the composite outcome (RR_c_: 0.74; 95% CI: 0.57 to 0.96) was lower than the effect on mortality (RR_d_: 0.56; 95% CI: 0.39 to 0.82), in patients with atrial fibrillation and coronary disease. Thus, the BACO index was 0.53 (95% CI: 0.21 to 0.86; p=0.0048).

The second study with statistically significant BACO was that by Lanz et al., which compared a self-expanding versus a balloon-expandable bioprosthesis for transcatheter aortic valve replacement in patients with symptomatic severe aortic stenosis [37]. The self-expanding bioprosthesis group presented an incidence of the composite outcome about 43% higher than observed in the balloon-expandable group (RR_c_: 1.43; 95% CI: 1.06 to 1.92). However, mortality in the first group was about three-times that of the second one (RR_d_: 2.96; 95% CI: 0.81 to 10.8). In this trial, the BACO index was 0.33 (95% CI: -0.1 to 0.76; p=0.002).

In the other study, Onland et al. evaluated the effect of systemic hydrocortisone compared with placebo on a composite outcome of death or bronchopulmonary dysplasia in very preterm infants [38]. The intervention was not associated with a significant change in the composite outcome incidence (RR_c_= 0.95; 95%CI: 0.84 to 1.08). However, the hydrocortisone group exhibited lower mortality compared to placebo (RR_d_: 0.65; 95% CI: 0.42 to 0.99). BACO index in this case was 0.11 (95% CI: -0.17 to 0.39; p<0.001).

Three other works were suggestive of a significant BACO, in which the RR of the composite outcome was closer to null value than the RR of mortality [33,34,39]. These studies included the clinical trials by Schuetz et al. (BACO index: 0.52; 95%CI: 0.07 to 0.98; p=0.04); Stone et al. (BACO index: 0.52; 95% CI: 0.07 to 0.98, p=0.03); and Nagel et al., which exhibited a negative point estimate of the BACO index (−0.06; 95% CI: -1.1 to 0.98, p=0.04).

In the positive controls, the CAPRICORN showed that carvedilol, compared with placebo, was not significantly associated with the composite outcome (RR = 0.94; 95% CI: 0.84 to 1.06) but was associated with a 22% reduction in mortality (BACO index: 0.24; 95% CI: 0.15 to 0.64). On the other hand, in the EXPEDITION study, the use of cariporide was associated with an 18% lower incidence of the composite outcome, but with a 53% higher mortality, compared with placebo (BACO index: -0.48; 95% CI: -1.03 to 0.08). The BACO indices of these two studies were significantly lower than one (p <0.001).

## 4. Discussion

Composite outcomes can make the interpretation of results challenging [1,8]. Differential effects on their less critical but more frequent components may result in a misleading impression about the impact of a treatment [2,45]. Therefore, it has been recommended that if there is a great variation between the effects on the components, the composite outcome should be abandoned [8].

However, assessing this is difficult, considering the asymmetric distribution of association measures and random variations. Some authors have evaluated the differences between the associations of both the composite outcome and mortality based on disagreement in statistical significance [1]. This type of comparison is biased because of the fatal outcome is a sub-element of the composite and will always have fewer events [4]. Therefore, there will be less power to evaluate an association with mortality.

The BACO index summarizes the relationship between the associations of the composite outcome and its most critical endpoint. Being based on the logarithms, the comparison is more consistent with the association measure distributions. On the other hand, the integration into a single index allows a unique statistical test for the null hypothesis.

Other authors stated they had planned to calculate a ratio between the efficacy for the composite outcome and that for mortality [7]. However, they considered that it would be problematic because the observations were not independent, as the death contributes to the composite. In that sense, the methodology proposed to obtain the BACO index solves this problem by considering overlapping observations [16,46,47].

Despite this, a limitation for an index based on the ratio of efficacy measures is that denominators close to zero lead to unstable or seemingly inflated results [7,48]. Hence, a BACO index should not be computed together with another whose reference effect is different. In other words, comparisons between BACO indices only make sense to contrast two or more composite outcomes when they consider the same reference value (e.g., of RR for mortality). In other circumstances, it is prudent to interpret the BACO indices only by classifying them into descriptive categories (overestimation, underestimation, and effect inversion) and considering the null hypothesis test.

In practice, the BACO index proved to be a simple measure to validate the composite outcome in clinical trials. Three of the analyzed studies had an index significantly lower than one, suggesting that the composite outcome underestimated the association between the intervention and the prognosis. Additionally, three other studies were suggestive of a similar trend, i.e., the composite outcomes seemed to dilute the associations that were stronger for mortality.

In the positive controls, the BACO index was significantly different from one. These studies have been well recognized as examples of bias associated with the composite outcome. After many discussions about the results of the CAPRICORN and EXPEDITION trials, their composite outcomes were disregard and the decisions based on the effects on all-cause mortality [9,10]. This would be consistent with the results of the BACO index.

Facing the need for objective guidelines, the BACO index may be a statistical tool to help interpret composite outcomes. Moreover, its application could be adapted according to the research context. For example, the level of significance could be adjusted depending on the desired sensitivity to identify a BACO. Furthermore, we can calculate the index from adjusted regression coefficients in observational studies that require controlling confusion.

This work is based on the expectation that the composite outcome must be in the same direction as its most relevant component. This is important when what is sought is that the composite outcome offers more events to increase power. In other cases, the composite outcome may not necessarily have a similar association magnitude as mortality. For example, when an intervention is expected to improve the quality of life without affecting total survival. However, even in these cases, a significant BACO could lead to reconsidering the necessity of a composite outcome or disaggregating the estimations for the component endpoints adequately.

## 5. Conclusions

The BACO index calculation could be incorporated into the analysis plan of clinical studies. Thus, based on a predefined rule, researchers could make impartial decisions about maintaining or replacing a composite outcome as the primary endpoint. Even if the researchers decide to base their conclusions on the composite outcome, a significant BACO should lead to caution that the association of the composite is stronger, weaker, or even opposite than that of its most critical component.

## Data Availability

All data necessary to verify the results were included in the manuscript or are available in the papers cited in the references.

## Acknowledgments

I thank my children, Alejandro and Ana Maria, for helping me organize the bibliographic material. I also thank Dr. Kai Arzheimer, for his guidance on Stata commands; and Alene Alder-Rangel, for her help with the English edition of the article.

## Appendix A. On the formula to estimate the variance of the BACO index

Let *φ_c_* and *φ_b_* be natural logarithms (*ln*) of the relative risks for the composite (*RR_c_*) and the target outcome (*RR_d_*), respectively. According the following contingency table:

**Table A.1.** Absolute frequency distribution of the composite outcome and its most critical component (target) in two treatment groups

|  |  |  |  |  |
| --- | --- | --- | --- | --- |
|  | Target outcome<br>(e.g. death) | Composite outcome |  |  |
|  |  | + | - | Total |
| Treatment group | + | $n_{t11}$ | 0 | $n_{t11}$ |
| | - | $n_{t21}$ | $n_{t22}$ | |
| | Total | $n_{t.1}$ | | $n_{t..}$ |
| Control group | + | $n_{c11}$ | 0 | $n_{c11}$ |
| | - | $n_{c21}$ | $n_{c22}$ | |
| | Total | $n_{c.1}$ | | $n_{c..}$ |

We have that:

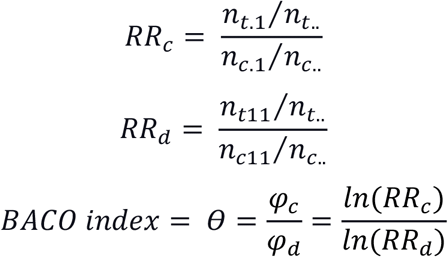

Consider the formula of the Taylor series estimator of the variance of an estimator of a ratio, *υ*(Ṙ), of random variables (Y and X, being Ṙ = Ẏ/Ẋ), according to which:

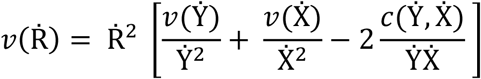

Where *υ*(Ẏ) and *υ*(Ẋ) are variances of the estimators of Y and X, respectively, and Ẏ, Ẋ is the covariance between them. Similarly, to calculate the variance BACO index,, we have:

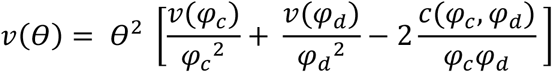

Where *υ*(*φ_c_*) and *υ*(*φ_d_*) are the variances of *φ_c_* and *φ_d_*, respectively, and *c*(*φ_c,_ φ_d_*), is their covariance.

Now, let a term *φ_c−d_* such as:

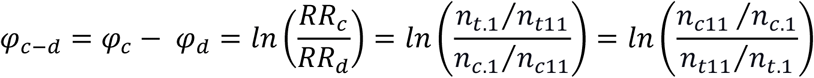

Consequently, variance estimators of *φ_c_*, *φ_d_*, and *φ_c−d_* would be respectively:

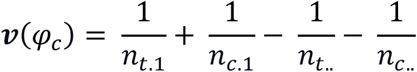

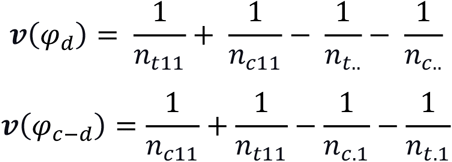

By considering the additive properties of variance and covariance, we have that

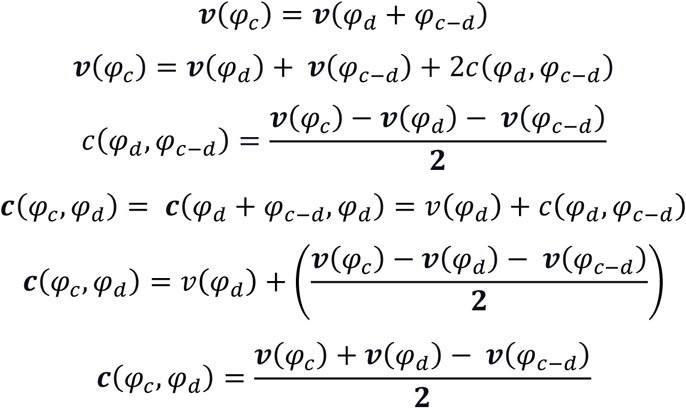

Moreover, because

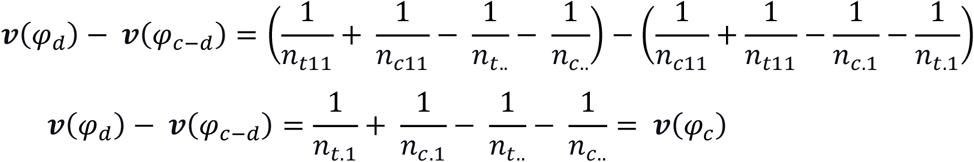

Then,

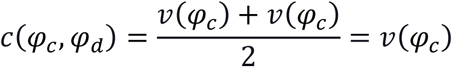

Consequently, the estimator of the variance of the BACO index can be

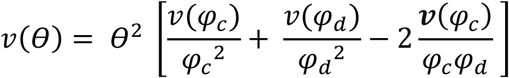

A 95% confidence interval for the BACO index is

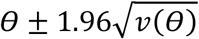

And the statistic *Z* with normal distribution to test the null hypothesis (BACO index = 1) is

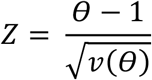

## Appendix B. Stata code to estimate and test the BACO index by combining regression parameters

Suppose we have a dataset with exposure or treatment variable “t”, composite outcome variable “c”, and the most critical endpoint variable “d” (i.e., death). For this example, take the data of the CAPRICORN study, which compared 975 patients allocated to experimental intervention vs. 984 patients in the control group [20]. The composite outcome occurred in 340 and 365 patients, including 116 and 151 deaths, respectively. Consequently, the RRs were 0.94 (95% CI: 0.84 – 1.06) for the composite outcome and 0.78 (95% CI: 0.62 – 0.97) for death. Before calculating the BACO index based on the relative risks, we should store and integrate the regression coefficients as described in the code that follows:

`poisson c t`

`est store coefcomp`

`poisson d t`

`est store coefdeath`

`suest coefcomp coefdeath`

In this example, the coefficients generated by the regression models were stored under the names “coefcomp” and “coefdeath”. Then, by using “suest” (seemingly unrelated estimation) command, we combine the stored parameter estimates and associated (co)variance matrices. Now we can use “nlcom” as described in the code that follows. The output (presented in the box) will include the estimate of the *BACO* index, its standard error, and a 95% confidence interval.

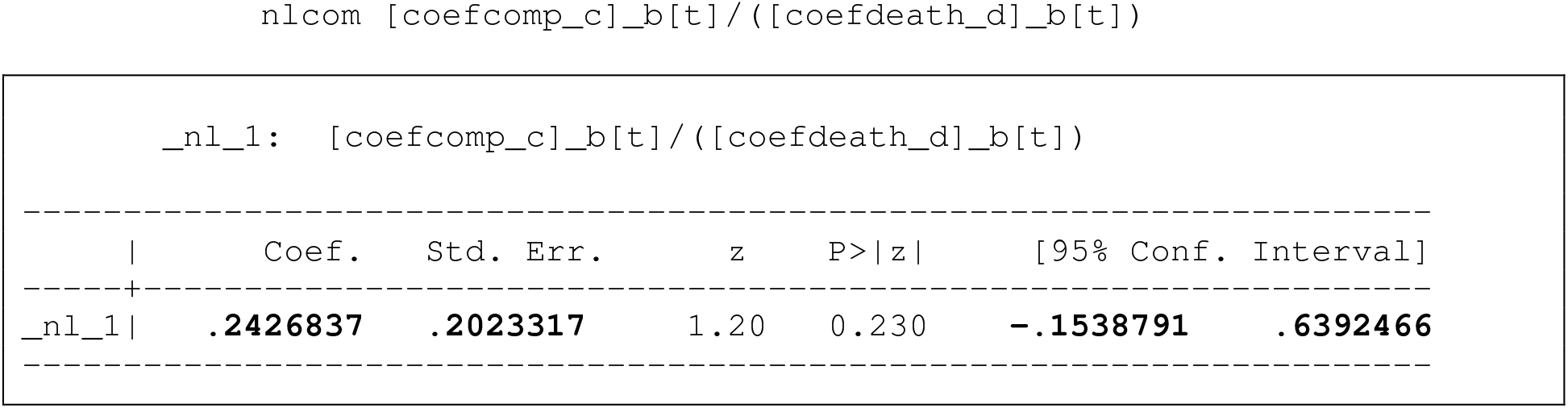

Please note that the previous p-value (0.23) refers to the null hypothesis of the BACO index equal to zero (therefore, it should not be considered). To calculate the p-value for the null hypothesis of the BACO index equals one, we can use the “testnl” command:

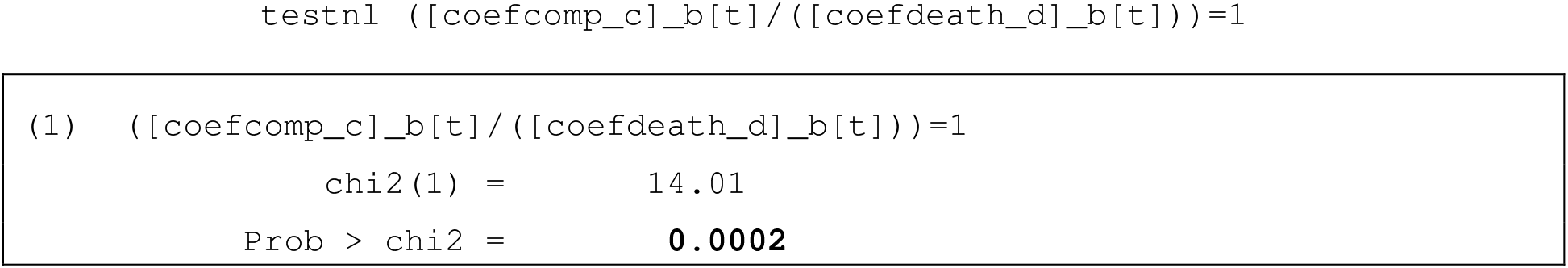

## Appendix C. Comparison of methods to calculate the standard error of the BACO index

Consider the four hypothetic composite outcomes described in the simulation (section 2.2). Table C.1 presents the standard error of each BACO index, 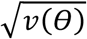, calculated both “directly” from the contingency tables, using formulas described in Appendix A, and by the nonlinear combination of regression parameters (NLCRP), using the tools described in Appendix B.

**Table C.1.** Standard error of BACO index calculated by two methods.

| Outcome | RR | BACO index | Standard Error |  |
| --- | --- | --- | --- | --- |
|  |  |  | Directly | NLCRP |
| Composite 1 (unbiased) | 0.6 | 1.00 | 0.2768 | 0.2769 |
| Composite 2 (overestimated effect) | 0.3 | 2.36 | 0.6222 | 0.6224 |
| Composite 3 (underestimated effect) | 0.8 | 0.44 | 0.1655 | 0.1655 |
| Composite 4 (inverted effect) | 2.0 | -1.36 | 0.5710 | 0.5712 |

In these examples, the two methods were virtually equivalent, and differences between them were observed only from the fourth decimal place (at the order of ten thousandths).

## References

[1] Freemantle N, Calvert M, Wood J, Eastaugh J, Griffin C. Composite Outcomes in Randomized Trials Greater Precision But With Greater Uncertainty? JAMA 2003;289:2554–9.

[2] Ferreira-González I, Busse JW, Heels-Ansdell D, Montori VM, Akl EA, Bryant DM, et al. Problems with use of composite end points in cardiovascular trials: Systematic review of randomised controlled trials. BMJ 2007;334:786–8. doi:10.1136/bmj.39136.682083.AE.

[3] Sankoh AJ, Li H, D’agostino RB. Use of composite endpoints in clinical trials. Stat Med 2014;33:4709–14. doi:10.1002/sim.6205.

[4] Pogue J, Thabane L, Devereaux P, Yusuf S. Testing for heterogeneity among the components of a binary composite outcome in a clinical trial. BMC Med Res Methodol 2010;10:49.

[5] Manja V, AlBashir S, Guyatt G. Criteria for use of composite end points for competing risks-a systematic survey of the literature with recommendations. J Clin Epidemiol 2017;82:4–11. doi:10.1016/j.jclinepi.2016.12.001.

[6] Armstrong PW, Westerhout CM. Composite End Points in Clinical Research. Circulation 2017;135:2299–307. doi:10.1161/CIRCULATIONAHA.117.026229.

[7] Cordoba G, Schwartz L, Woloshin S, Bae H, Gøtzsche PC. Definition, reporting, and interpretation of composite outcomes in clinical trials: systematic review. BMJ 2010;341:c3920. doi:10.1136/bmj.c3920.

[8] Montori VM, Permanyer-Miralda G, Ferreira-González I, Busse JW, Pacheco-Huergo V, Bryant D, et al. Validity of composite end points in clinical trials. BMJ 2005;330:594–6. doi:10.1136/bmj.330.7491.594.

[9] Pocock SJ, Stone GW. The Primary Outcome Fails — What Next? N Engl J Med 2016;375:861–70. doi:10.1056/NEJMra1510064.

[10] Pocock SJ, Stone GW. The Primary Outcome Is Positive — Is That Good Enough? N Engl J Med 2016;375:971–9. doi:10.1056/NEJMra1601511.

[11] Pogue J, Devereaux PJ, Thabane L, Yusuf S. Designing and Analyzing Clinical Trials with Composite Outcomes: Consideration of Possible Treatment Differences between the Individual Outcomes. PLoS One 2012;7:34785. doi:10.1371/journal.pone.0034785.

[12] Gelijns AC. Modern Methods of Clinical Investigation. Washington (DC): NATIONAL ACADEMY PRESS; 1990. doi:10.17226/1550.

[13] Wolter KM. Taylor Series Methods. In: Wolter KM, editor. Introd. to Var. Estim., Springer New York; 2007, p. 226–71. doi:10.1007/978-0-387-35099-8_6.

[14] Carter RE, Zhang X, Woolson RF, Apfel CC. Statistical Analysis of Correlated Relative Risks. J Data Sci 2009;7:397–407.

[15] Barros AJD, Hirakata VN. Alternatives for logistic regression in cross-sectional studies: an empirical comparison of models that directly estimate the prevalence ratio. BMC Med Res Methodol 2003;3:21. doi:10.1186/1471-2288-3-21.

[16] Stata Press. STATA BASE REFERENCE MANUAL Release 15. 15th ed. Texas: Stata Press; 2017.

[17] Popma JJ, Michael Deeb G, Yakubov SJ, Mumtaz M, Gada H, O’Hair D, et al. Transcatheter aortic-valve replacement with a self-expanding valve in low-risk patients. N Engl J Med 2019;380:1706–15. doi:10.1056/NEJMoa1816885.

[18] Benjamin DJ, Berger JO, Johannesson M, Nosek BA, Wagenmakers E-J, Berk R, et al. Redefine statistical significance. Nat Hum Behav 2018;2:6–10. doi:10.1038/s41562-017-0189-z.

[19] Benjamin DJ, Berger JO. Three Recommendations for Improving the Use of *p* -Values. Am Stat 2019;73:186–91. doi:10.1080/00031305.2018.1543135.

[20] Dargie HJ. Effect of carvedilol on outcome after myocardial infarction in patients with left-ventricular dysfunction: The CAPRICORN randomised trial. Lancet 2001;357:1385–90. doi:10.1016/S0140-6736(00)04560-8.

[21] Mentzer RM, Bartels C, Bolli R, Boyce S, Buckberg GD, Chaitman B, et al. Sodium-Hydrogen Exchange Inhibition by Cariporide to Reduce the Risk of Ischemic Cardiac Events in Patients Undergoing Coronary Artery Bypass Grafting: Results of the EXPEDITION Study. Ann Thorac Surg 2008;85:1261–70. doi:10.1016/j.athoracsur.2007.10.054.

[22] Holm NR, Mäkikallio T, Lindsay MM, Spence MS, Erglis A, Menown IBA, et al. Percutaneous coronary angioplasty versus coronary artery bypass grafting in the treatment of unprotected left main stenosis: updated 5-year outcomes from the randomised, non-inferiority NOBLE trial. Lancet 2019. doi:10.1016/S0140-6736(19)32972-1.

[23] Katheria A, Reister F, Essers J, Mendler M, Hummler H, Subramaniam A, et al. Association of umbilical cord milking vs delayed umbilical cord clamping with death or severe intraventricular hemorrhage among preterm infants. JAMA -J Am Med Assoc 2019;322:1877–86. doi:10.1001/jama.2019.16004.

[24] Schüpke S, Neumann F-J, Menichelli M, Mayer K, Bernlochner I, Wöhrle J, et al. Ticagrelor or Prasugrel in Patients with Acute Coronary Syndromes. N Engl J Med 2019;381:1524–34. doi:10.1056/NEJMoa1908973.

[25] Brott TG, Calvet D, Howard G, Gregson J, Algra A, Becquemin JP, et al. Long-term outcomes of stenting and endarterectomy for symptomatic carotid stenosis: a preplanned pooled analysis of individual patient data. Lancet Neurol 2019;18:348–56. doi:10.1016/S1474-4422(19)30028-6.

[26] Tomaniak M, Chichareon P, Onuma Y, Deliargyris EN, Takahashi K, Kogame N, et al. Benefit and risks of aspirin in addition to ticagrelor in acute coronary syndromes: A post hoc analysis of the randomized global leaders trial. JAMA Cardiol 2019;4:1092–101. doi:10.1001/jamacardio.2019.3355.

[27] Wu Y, Li S, Patel A, Li X, Du X, Wu T, et al. Effect of a Quality of Care Improvement Initiative in Patients with Acute Coronary Syndrome in Resource-Constrained Hospitals in China: A Randomized Clinical Trial. JAMA Cardiol 2019;4:418–27. doi:10.1001/jamacardio.2019.0897.

[28] Hahn JY, Song Y Bin, Oh JH, Chun WJ, Park YH, Jang WJ, et al. Effect of P2Y12 Inhibitor Monotherapy vs Dual Antiplatelet Therapy on Cardiovascular Events in Patients Undergoing Percutaneous Coronary Intervention: The SMART-CHOICE Randomized Clinical Trial. JAMA -J Am Med Assoc 2019;321:2428–37. doi:10.1001/jama.2019.8146.

[29] Packer DL, Mark DB, Robb RA, Monahan KH, Bahnson TD, Poole JE, et al. Effect of Catheter Ablation vs Antiarrhythmic Drug Therapy on Mortality, Stroke, Bleeding, and Cardiac Arrest among Patients with Atrial Fibrillation: The CABANA Randomized Clinical Trial. JAMA -J Am Med Assoc 2019;321:1261–74. doi:10.1001/jama.2019.0693.

[30] Macdougall IC, White C, Anker SD, Bhandari S, Farrington K, Kalra PA, et al. Intravenous Iron in Patients Undergoing Maintenance Hemodialysis. N Engl J Med 2019;380:447–58. doi:10.1056/NEJMoa1810742.

[31] Mack MJ, Leon MB, Thourani VH, Makkar R, Kodali SK, Russo M, et al. Transcatheter Aortic-Valve Replacement with a Balloon-Expandable Valve in Low-Risk Patients. N Engl J Med 2019;380:1695–705. doi:10.1056/NEJMoa1814052.

[32] Yasuda S, Kaikita K, Akao M, Ako J, Matoba T, Nakamura M, et al. Antithrombotic Therapy for Atrial Fibrillation with Stable Coronary Disease. N Engl J Med 2019;381:1103–13. doi:10.1056/NEJMoa1904143.

[33] Schuetz P, Fehr R, Baechli V, Geiser M, Deiss M, Gomes F, et al. Individualised nutritional support in medical inpatients at nutritional risk: a randomised clinical trial. Lancet (London, England) 2019;393:2312–21. doi:10.1016/S0140-6736(18)32776-4.

[34] Stone GW, Kappetein AP, Sabik JF, Pocock SJ, Morice M-C, Puskas J, et al. Five-Year Outcomes after PCI or CABG for Left Main Coronary Disease. N Engl J Med 2019;381:1820–30. doi:10.1056/NEJMoa1909406.

[35] Zenati MA, Bhatt DL, Bakaeen FG, Stock EM, Biswas K, Gaziano JM, et al. Randomized Trial of Endoscopic or Open Vein-Graft Harvesting for Coronary-Artery Bypass. N Engl J Med 2019;380:132–41. doi:10.1056/NEJMoa1812390.

[36] Iversen K, Ihlemann N, Gill SU, Madsen T, Elming H, Jensen KT, et al. Partial Oral versus Intravenous Antibiotic Treatment of Endocarditis. N Engl J Med 2019;380:415–24. doi:10.1056/NEJMoa1808312.

[37] Lanz J, Kim WK, Walther T, Burgdorf C, Möllmann H, Linke A, et al. Safety and efficacy of a self-expanding versus a balloon-expandable bioprosthesis for transcatheter aortic valve replacement in patients with symptomatic severe aortic stenosis: a randomised non-inferiority trial. Lancet 2019;394:1619–28. doi:10.1016/S0140-6736(19)32220-2.

[38] Onland W, Cools F, Kroon A, Rademaker K, Merkus MP, Dijk PH, et al. Effect of Hydrocortisone Therapy Initiated 7 to 14 Days after Birth on Mortality or Bronchopulmonary Dysplasia among Very Preterm Infants Receiving Mechanical Ventilation: A Randomized Clinical Trial. JAMA - J Am Med Assoc 2019;321:354–63. doi:10.1001/jama.2018.21443.

[39] Nagel E, Greenwood JP, McCann GP, Bettencourt N, Shah AM, Hussain ST, et al. Magnetic Resonance Perfusion or Fractional Flow Reserve in Coronary Disease. N Engl J Med 2019;380:2418–28. doi:10.1056/NEJMoa1716734.

[40] Ho KM, Rao S, Honeybul S, Zellweger R, Wibrow B, Lipman J, et al. A Multicenter Trial of Vena Cava Filters in Severely Injured Patients. N Engl J Med 2019;381:328–37. doi:10.1056/NEJMoa1806515.

[41] Van Spall HGC, Lee SF, Xie F, Oz UE, Perez R, Mitoff PR, et al. Effect of Patient-Centered Transitional Care Services on Clinical Outcomes in Patients Hospitalized for Heart Failure: The PACT-HF Randomized Clinical Trial. JAMA 2019;321:753–61. doi:10.1001/jama.2019.0710.

[42] Kozhuharov N, Goudev A, Flores D, Maeder MT, Walter J, Shrestha S, et al. Effect of a Strategy of Comprehensive Vasodilation vs Usual Care on Mortality and Heart Failure Rehospitalization Among Patients With Acute Heart Failure: The GALACTIC Randomized Clinical Trial. JAMA 2019;322:2292–302. doi:10.1001/jama.2019.18598.

[43] Nguyen HQ, Moy ML, Liu I-LA, Fan VS, Gould MK, Desai SA, et al. Effect of Physical Activity Coaching on Acute Care and Survival Among Patients With Chronic Obstructive Pulmonary Disease: A Pragmatic Randomized Clinical Trial. JAMA Netw Open 2019;2:e199657. doi:10.1001/jamanetworkopen.2019.9657.

[44] Vousden N, Lawley E, Nathan HL, Seed PT, Gidiri MF, Goudar S, et al. Effect of a novel vital sign device on maternal mortality and morbidity in low-resource settings: a pragmatic, stepped-wedge, cluster-randomised controlled trial. Lancet Glob Heal 2019;7:e347–56. doi:10.1016/S2214-109X(18)30526-6.

[45] Lim E, Brown A, Helmy A, Mussa S, Altman DG. Composite outcomes in cardiovascular research: A survey of randomized trials. Ann Intern Med 2008;149:612–7. doi:10.7326/0003-4819-149-9-200811040-00004.

[46] Oehlert GW. A Note on the Delta Method. Am Stat 1992;46:27–9. doi:10.1080/00031305.1992.10475842.

[47] Arzheimer K. nlcom and the Delta Method 2012. https://www.kai-arzheimer.com/delta-method-nlcom/amp/ (accessed January 27, 2020).

[48] Krogsbøll LT, Hróbjartsson A, Gøtzsche PC. Spontaneous improvement in randomised clinical trials: metaanalysis of three-armed trials comparing no treatment, placebo and active intervention. BMC Med Res Methodol 2009;9:1. doi:10.1186/1471-2288-9-1.

